# Common and separable neural alterations in adult and adolescent depression – evidence from neuroimaging meta-analyses

**DOI:** 10.1101/2024.02.27.24303428

**Authors:** Mercy Chepngetich Bore, Xiqin Liu, Keith M Kendrick, Bo Zhou, Jie Zhang, Benjamin Klugah-Brown, Benjamin Becker

**Author notes:** Corresponding authors Benjamin Becker Email address Postal/mail address: University of Electronic Science and Technology of China, Xiyuan Ave 2006, 611731 Chengdu, China. University of Hong Kong Pokfulam, Hong Kong, China Benjamin Klugah-Brown Email address Postal/mail address: University of Electronic Science and Technology of China, Xiyuan Ave 2006, 611731 Chengdu, China.

## Abstract

Depression is a highly prevalent and debilitating disorder that often begins in adolescence. However, it remains unclear whether adults and adolescents with depression exhibit common or separate brain dysfunctions during reward processing. We aimed to identify common and separable neurofunctional alterations during receipt of rewards and brain structure in adolescents and adults with depression. A coordinate-based meta-analysis was employed using Seed-based d mapping with permutation of subject images (SDM-PSI). Compared with healthy controls, both age groups exhibited common activity decreases in the right striatum (putamen, caudate) and subgenual ACC. Adults with depression showed decreased reactivity in the right putamen and subgenual ACC, while adolescents with depression showed decreased activity in the left mid cingulate, right caudate but increased reactivity in the right post central gyrus. This meta-analysis revealed shared (caudate) and separable (putamen and mid cingulate cortex) reward-related alterations in adults and adolescents with depression. The findings suggest age-specific neurofunctional alterations and stress the importance of adolescent-specific interventions that target social functions.

## 1. Introduction

Adolescence is a critical period for the emergence of major depressive disorder (MDD). During this developmental stage, majority of cases will experience the onset of this highly prevalent and debilitating mental disorder (Kessler et al., 2001; Rice et al., 2019). With over 300 million people affected worldwide, depression represents a leading cause of morbidity and disability and is accompanied by a strongly increased risk of suicide (Friedrich, 2017; Hawton et al., 2013). On the symptomatic level, MDD is primarily characterized by persistent depressed mood and anhedonia (loss of interest of previously rewarding events) (Husain and Roiser, 2018; Luking et al., 2016b; Malhi and Mann, 2018; Treadway and Zald, 2011).

Mounting evidence suggests that dysfunctional reward processing represents a candidate mechanism underlying the development and maintenance of depressed mood and anhedonia (Bore et al., 2023; Husain and Roiser, 2018). Rewards are described as events that elicit pleasurable or positive affective experiences (White, 2011) and the pursuit of rewards is crucial for survival because it is one of the strongest motivators in both animals and humans (Sosa and Giocomo, 2021). Adaptative reward processing is crucial for hedonic experience, learning from rewards and shaping future motivational behavior (Berridge and Robinson, 2003; Rizvi et al., 2016; Yang et al., 2021; Zhou et al., 2023) and dysregulations in this domain form the core of the Research Domain Criteria (RDoC) framework for positive valence systems and may represent a candidate mechanism underpinning anhedonia (Insel et al., 2010; Oldham et al., 2018).

Dysregulations in this domain form the core of the Research Domain Criteria (RDoC) framework for positive valence systems and may represent a candidate mechanism underpinning anhedonia (Insel et al., 2010; Oldham et al., 2018). The neural basis of reward processing has been extensively mapped across species and has been closely linked to mesocortico-limbic pathways, in particular the (ventral) striatum and its connection with prefrontal and limbic regions (Mas-Herrero et al., 2021; Schultz et al., 2000; Xu et al., 2023; Zhou et al., 2023). While mounting evidence from functional MRI studies indicates that reward processing dysfunctions in adult patients with mental disorders, including those with depression map onto these circuitries (Bore et al., 2023; Klugah-Brown et al., 2020; Zeng et al., 2022; Zimmermann et al., 2019; Zimmermann et al., 2018), less is known about the neural basis of reward processing dysfunctions in adolescent depression and whether these resemble those observed in adults.

Although depression is presented similarly in many aspects among adults and adolescents, it is well documented that these two groups have important etiological differences and furthermore, they exhibit distinct physical and emotional manifestations as well as responses to life stressors (Rice et al., 2019). The adolescence period tends to herald a myriad of physical, emotional and psychological changes that increase the risk of onset and recurrent episodes of depressive symptoms which often evolve into the full picture of MDD (Davey et al., 2008; Dunn and Goodyer, 2018).

Adolescence is period of significant physical, emotional and psychological changes that are accompanied by profound developmental transformations. During this period, adolescents tend to engage in more reward-seeking behaviors as well as experience rewards more intensely compared to other age-groups (Ernst et al., 2005). This level of reward-dependence during adolescence may be associated with the high prevalence rates of first episodes of depression (Forbes and Dahl, 2012). On the clinical level, important differences exist in the manifestation of adolescent versus adult depression. While adult MDD is primarily characterized by persistent depressive mood and anhedonia adolescent depression is mostly characterized by vegetative symptoms including fatigue, insomnia, appetite and weight disturbance (Jiang et al., 2023).

Furthermore, somatic profiles in clinically depressed children and adolescents show occurrence of symptoms such as persistent headaches and muscle pain (Nardi et al., 2013). Moreover, some studies have shown that adolescents with depression often experience remission after antidepressant treatment (McMakin et al., 2012) while adults commonly respond poorly to antidepressant medication (Uher et al., 2011). Understanding common and distinct reward-related neural dysfunctions in adolescents and adult depression may thus contribute to determine more precise therapeutic interventions in depression.

The neural process of receiving rewards has been consistently found impaired in patients with depression compared to healthy controls, in regions such as the ventral striatum, orbitofrontal/ventromedial prefrontal cortex, posterior cingulate and amygdala (Oldham et al., 2018; Pizzagalli et al., 2009; Rizvi et al., 2016; Segarra et al., 2016). While previous meta-analyses of fMRI case control studies documented robust neurofunctional reward alterations in adult (Keren et al., 2018; Pizzagalli et al., 2009) or adolescent depression (Luking et al., 2016a; McCabe et al., 2012; Stringaris et al., 2015) a direct comparison of the alterations is lacking.

Convergent evidence suggests that adults with depression exhibit aberrant activity in the striatum and frontal cortex (Geugies et al., 2019; Keren et al., 2018; Ng et al., 2019; Russo and Nestler, 2013; Solomonov et al., 2023), mostly consistent decreased striatal response during rewad receipt (Oh et al., 2021; Smoski et al., 2009). While adolescents with depression have shown attenuated responses during reward outcome (Luking et al., 2016a; McCabe et al., 2012), it remains unclear whether the findings converge on common neurofunctional alterations across the age groups and whether separable regions may underpin the distinct symptomatic profiles in adult versus adolescent depression.

To this end, the present pre-registered systematic neuroimaging meta-analysis aimed to determine common and separable neurofunctional alterations during reward processing in adolescents and adults. Meta-analyses have in the recent past been utilized to systematically and quantitatively synthesize and compare findings of previously published brain imaging studies and may allow to overcome heterogeneity, generalizability and consistency limitations (Gurevitch et al., 2018; Kotov et al., 2010; Zhou et al., 2022). The present meta-analysis aimed to determine common and distinct brain functional alterations during reward processing in adults and adolescents with depression. Specifically, we aim to determine (a) general reward alterations in both adults and adolescents with depression, (b) robust neurofunctional alterations during reward processing in adolescents with depression, (c) meta-analytically characterize the identified regions on the behavioral, molecular and network level, (d) explore whether commonly reported brain structural alterations in depression overlap with the identified functional alterations (Liu et al., 2022). We hypothesized that adults would exhibit reward-processing alterations in the striatum while adolescents would potentially exhibit striatal dysfunction as well as alterations in other regions involved in value and cognitive control processing or those involved in social and self-referential processing such as the default mode network. The analyses were pre-registered (https://osf.io/wc8eh) and implemented in Seed-based d Mapping with Permutation of Subject Images (SDM-PSI) version 6.21 (https://www.sdmproject.com/), a novel and well established technique for performing neuroimaging meta-analyses (Albajes-Eizagirre et al., 2019a).

## 2. Methods

### 2.1. Search strategy and selection criteria

The current meta-analysis follows widely accepted procedures for conducting coordinate-based meta-analyses (Müller et al., 2018) and the Preferred Reporting Items for Systematic Reviews and Meta-Analyses (PRISMA) guidelines (Page et al., 2021). An extensive literature search was conducted on three databases; Pubmed (https://www.ncbi.nlm.nih.gov/pubmed), Web of Science (https://www.webofscience.com/wos/alldb/basic-search) and, PsycInfo https://www.apa.org/pubs/databases/psycinfo/). To increase the credibility and interpretability of the results, the current meta-analysis was pre-registered in the Open Science Framework (OSF) repository prior to data analysis (https://osf.io/wc8eh). Studies included in the main reward outcome analysis of adults and adolescents with depression are listed (Admon et al., 2015; Canli et al., 2004; Chantiluke et al., 2012; Davey et al., 2011; Dichter et al., 2012; Epstein et al., 2006; Fischer et al., 2019; Forbes et al., 2009; Fournier et al., 2013; Gotlib et al., 2010; Gotlib et al., 2005; Gradin et al., 2015; Hall et al., 2014; Harlé et al., 2023; Keedwell et al., 2005; Knutson et al., 2008; Kumari et al., 2003; Luking et al., 2016a; Martin-Soelch et al., 2021; McCabe et al., 2009; Morgan et al., 2019; Oh et al., 2021; Olino et al., 2015; Pizzagalli et al., 2009; Segarra et al., 2016; Sharp et al., 2014; Smoski et al., 2009; Smoski et al., 2011; Wiggins et al., 2017; Yoon et al., 2022) (see **Supplementary Table2**).

### 2.2. Coordinate-based meta-analytic approach

The coordinate-based meta-analysis was performed using SDM-PSI toolbox (https://www.sdmproject.com). Peak coordinates of between-group differences (patients versus healthy subjects) and their corresponding effect sizes (t/z values) were extracted. z-values were converted to t-values using the standard statistical converter (https://www.sdmproject.com/utilities/?show=Coordinates). (b) Preprocessing was performed to estimate the lower and upper bounds (Hedges’ g) of the most probable effect size images (Radua et al., 2014). (c) The mean was calculated using maximum-likelihood estimation (MLE) and meta-analysis of non-statistically significant unreported effects (MetaNSUE) algorithms. This step estimates the most likely effect sizes generating several imputations according to estimates within the bounds (Albajes-Eizagirre et al., 2019b). (d) Imputed study images are then recreated. (e) Finally, the permutation test evaluated the combined meta-analysis images for statistical significance (Albajes-Eizagirre et al., 2019c).

Visualization of brain maps was performed using MRIcroGL (NITRC: MRIcroGL: Tool/Resource Info). All analyses were considered significant at p < .0025 uncorrected, with a threshold of k=10 voxels. This threshold balances Type I and Type II errors, as shown in a previous meta-analysis (Liu et al., 2022). Three main meta-analysis, including all adult and adolescent studies (n=30), adult studies (n=18) and adolescent studies (n=12) were conducted. First, a general meta-analysis of all reward outcome studies of adults and adolescents with depression was performed. Next, a meta-analysis of all reward outcome studies on adults with depression was conducted. Finally, a meta-analysis of all fMRI studies on adolescents with depression was conducted. Subsequent analyses mostly compared the peak regions identified in the meta-analysis results of the two groups.

### 2.3. Selection criteria

Original fMRI studies of adults and adolescents with depression were screened, and suitable studies were included according to the pre-defined inclusion and exclusion criteria. Additionally, relevant articles from other sources, such as review articles, were included. The selection criteria encompassed studies written in English and published between January 2001 and January 2023, reporting whole brain results in standard stereotactic space (Talairach or Montreal Neurological Institute). To augment the sample size of the adolescent group, one study was included outside the pre-registered inclusion period (Harlé et al., 2023). The data extraction process, involving details such as authors’ names, paper titles, publication years, age and gender of participants, medication status, experimental paradigms, etc., was carried out by M.C.B and B.K.B. Any discrepancies during the literature search, screening, or inclusion process were resolved through consensus with B.B. The literature search adopted a three-step approach, covering adult and adolescent fMRI studies, as well as exploratory VBM studies. For adult and adolescent fMRI studies, the primary contrast focused on the receipt of rewards in both age groups. The search terms included: “functional magnetic resonance imaging OR fMRI” AND “depression OR major depressive disorder OR unipolar depression OR sub-clinical depression OR at-risk of depression” combined with either “monetary reward” OR “monetary incentive delay” OR “natural reward” OR “social reward”. For adolescent depression studies, the search terms were: “functional magnetic resonance imaging” OR “fMRI” AND “depression” OR “major depressive disorder” AND “reward”. Finally, the exploratory VBM dataset search terms included: “Depression” AND “voxel-based morphometry”, OR (“VBM”), OR (“gray matter”). The exclusion criteria included: (a) studies on geriatric (elderly) depression (participants > 60 years old), (b)studies centered on other mood disorders, e.g., bipolar disorder and postpartum depression, (b) studies on region-of-interest (ROI) results only, (c) studies of other reward processes e.g., reward learning, (d) studies reporting results from already included studies.

### 2.4. Network, behavioral, genetic and receptor-level analyses

Firstly, the main meta-analysis aimed at determining common and distinct reward alterations in adult and adolescent depression. Network analyses were performed using the peak coordinates of the identified regions representing nodes of separable networks. The identified signatures were subjected to network analyses, with peak coordinates of the identified regions defining nodes in separate networks. Seed-to-whole-brain functional connectivity analyses from an independent dataset were utilized to provide a more fine-grained mapping of the common and separable intrinsic network organization of the identified brain regions. Data collection involved 100 subjects from the Human Connectome Project. Preprocessing was performed using Data Processing Assistant for Resting-State fMRI (http://rfmri.org/DPARSF) with the following steps: (a) Removal of the first 5 volumes of the time-series, (b) Motion correction using rigid body rotation and translation, while excluding subjects with a maximum motion > 2 mm or 2°, (c) Normalization into standard stereotactic space, which included co-registration to and the matching segmentation matrix was applied to the functional time series. Spheres of 6mm radius centered at the corresponding peak coordinates (caudate: 12, 6, -8, putamen: 18, 4, -8 and mid cingulate cortex: 2, -28, 38) served as seed regions for the voxel-wise analyses completed in Data Processing Assistant for Resting-State fMRI (http://rfmri.org/DPARSF).

Secondly, distinct behavioral functions of the identified signatures were identified using the Brain Annotation Toolbox (BAT) (Liu et al., 2019). The top ten functional characterizations of each region were extracted and plotted according to their p-values, starting with the most significant (p_perm_ < 0.05 was considered significant). Thirdly, to investigate the separable genetic underpinnings of the identified brain regions, gene expressions were determined using the BAT toolbox. This analysis identified genes with the highest expressions in the identified regions. The functions of the top ten genes were further determined using PubMed (https://www.ncbi.nlm.nih.gov/gene).

Finally, neurotransmitter analysis of dopamine (DA) and serotonin (5HT) were performed to compare their mean density values in the identified regions. Dopamine and serotonin were selected on the basis of their role in motivation and reward processing dysregulations in mental disorders such as depression (Liu et al., 2019; Martins et al., 2021). They steps included: a) Acquisition of Positron Emission Tomography (PET) atlases of Serotonin (5-HT1B) (https://xtra.nru.dk/FS5ht-atlas/) and Dopamine transporter (DA) (https://www.nitrc.org/projects/spmtemplates) maps. Refence regions will be removed using WFU PickAtlas (v2.4) i.e., occipital and cerebellum regions of DA and 5-HT1B maps. Spherical ROIs (6mm) were thereafter built using Marsbar ROI toolbox (http://marsbar.sourceforge.net) in the identified regions. Mean densities of the neurotransmitters were acquired using the Cognitive and Affective Neuroscience Lab (CANlab) Core toolbox (https://canlabweb.colorado.edu/).

### 2.5. Tests for heterogeneity, sensitivity and publication bias

Heterogeneity, sensitivity and publication bias tests were performed according to standard procedures. Tests for heterogeneity are important when considering the studies included in the meta-analysis used the same experimental protocols and procedures. Between-study heterogeneity for all significant clusters of the main meta-analyses was examined by the *I^2^* index (represents a proportion of the total variation in heterogeneity derived from studies) (Higgins and Thompson, 2002). *I^2^*scores can indicate low (>25%), moderate (>50%) and high (>70%) heterogeneity (Martins et al., 2021). Jackknife sensitivity analysis (repeating the meta-analysis excluding one study every time to show that findings were not driven by inclusion of some studies) was conducted on Anisotropic Effect Size-Signed Differential Mapping (AES-SDM) to measure the robustness of the meta-analysis findings. Publication bias was statistically evaluated and interpreted by Egger’s tests and funnel plots (an asymmetric plot and p<0.05 were recognized as significant).

### 2.6. Exploratory analyses – meta-regression and linear model analysis

An exploratory VBM meta-analysis of adult and adolescent depression was performed to explore whether structural alterations overlap with the functional alterations determined in depression (Liu et al., 2022). Confounding effects of medication in the adult group was assessed by the linear model function of SDM-PSI. Here, medication and non-medication studies were coded as 1 and 0, respectively. We further performed a sub-group meta-analysis including only unmedicated studies to validate the stability of the results. Moreover, the influence of potential confounding effects of age and gender (number of females) were examined by meta-regression.

## 3. Results

### 3.1. Demographic and clinical data summary of all included studies

The extensive literature search produced 75 suitable fMRI and VBM studies on adult and adolescent depression, i.e., 18 fMRI studies on adult depression, 12 fMRI studies on adolescent depression, 7 adolescent VBM studies and 44 adult VBM studies on depression. A total of n= 1,492 participants (n=734 patients, n=758 healthy controls) were included in the main fMRI meta-analysis. The database included patient data from adult studies (*n*=410, mean age=34.39, *SD*=9.61) and adolescent studies (*n*=324, mean age=13.29, *SD*=1.56). Healthy controls data from adult studies (*n*=423, mean age=32.23, *SD*=8.72) and adolescent studies (*n*=335, mean age=13.52, *SD*=1.72). There were no significant group differences between patients and healthy controls in the adult (p=0.32, t=0.99) and adolescent (p=0.91, t=-0.11) groups. The exploratory VBM meta-analysis included a total of 44 studies (7 adolescent studies and 37 adult studies). **Fig. 1** shows the flow diagram of the selection criteria. **Fig. 2** shows the workflow of all the main analyses that were conducted. A summary of all included studies is presented in **Supplementary Table 1.**

**Fig. 1.**
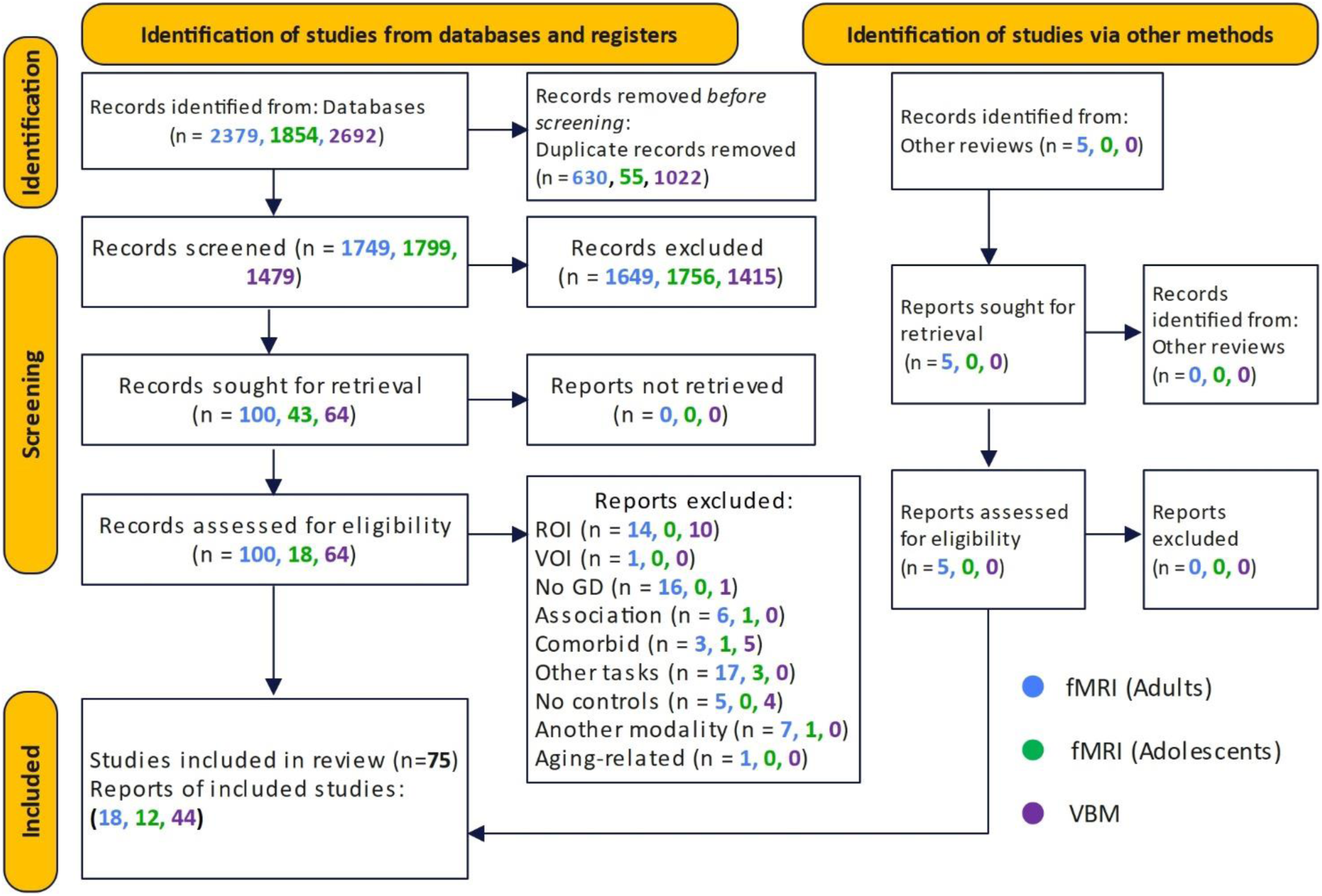
PRISMA flow diagram of the selection process

**Fig. 2.**
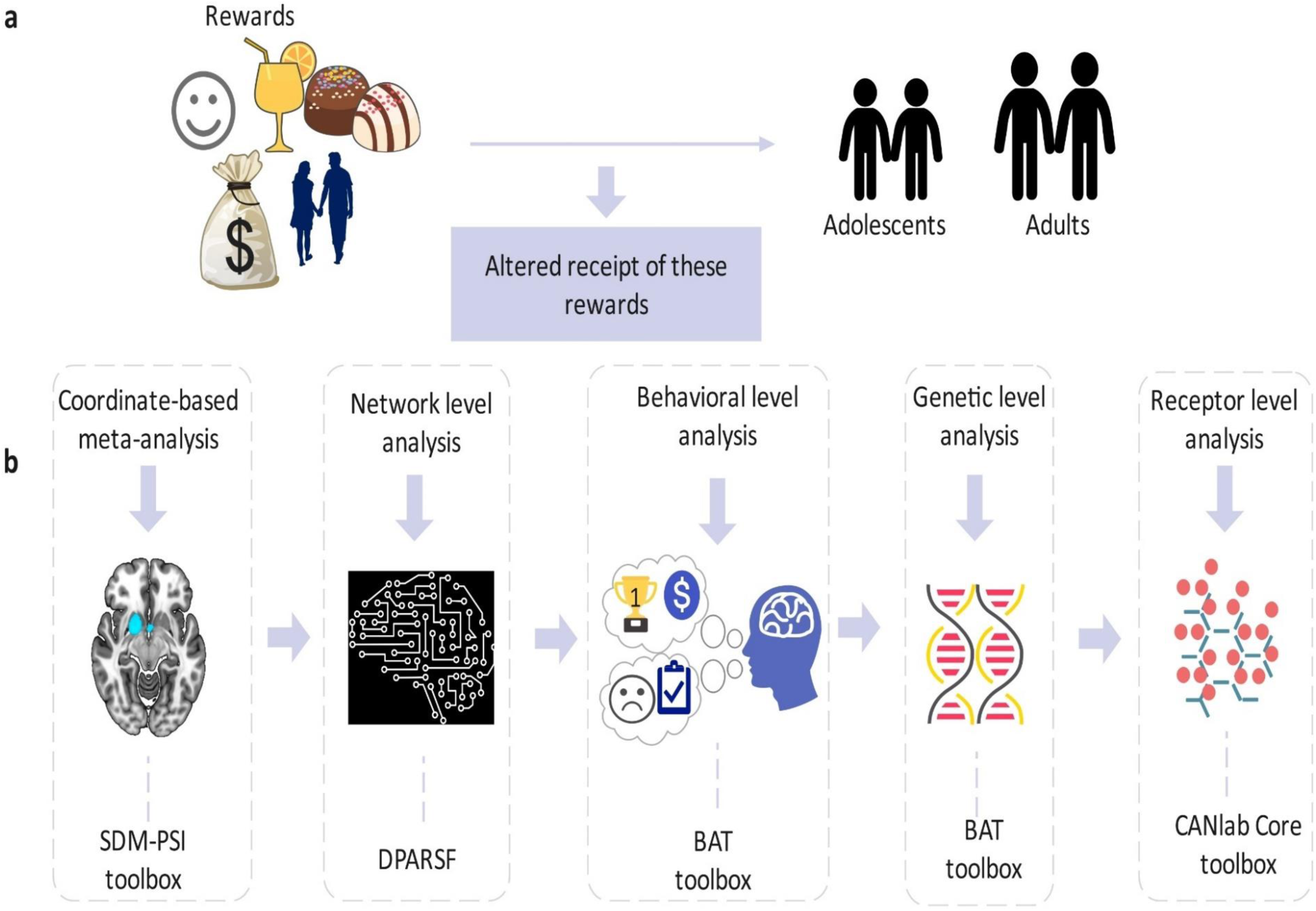
**a** General illustration of altered receipt of rewards in adolescents and adults with depression. **b** Work flow of main analyses in the current study.

### 3.2. Meta-analytic results of reward dysfunctions across all fMRI studies

The meta-analysis of all pooled studies of adult and adolescent depression during receipt of rewards revealed decreased activation in the subgenual anterior cingulate and right striatum (putamen extending to the caudate) (**Fig. 3a**). Compared to healthy controls, adults with depression showed decreased activation in the right putamen and the subgenual anterior cingulate cortex (**Fig. 3b**). However, adolescents with depression showed decreased activation in the left median cingulate, right caudate nucleus and inter-hemispheric regions compared to healthy controls (**Fig. 3c**) However, there was one peak of increase in the right postcentral gyrus in adolescents with depression versus healthy controls. Clusters were considered significant at p < .0025 uncorrected across all meta-analyses. A summary of all significant clusters and their effect sizes are presented on **Table 1**.

**Fig. 3.**
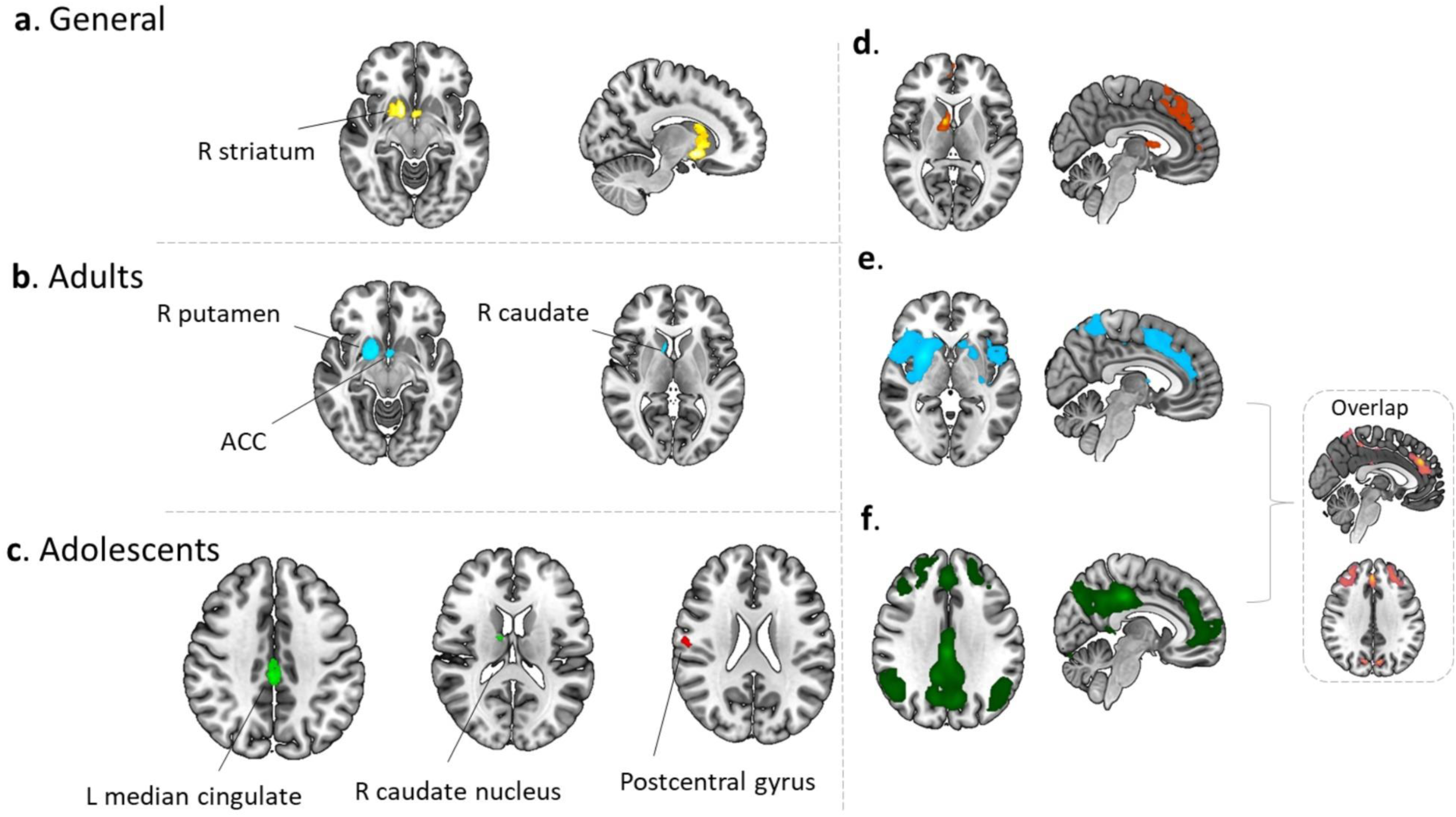
Results of the main meta-analyses of altered reward processing in adolescent and adult depression compared to healthy controls. **a** Overall meta-analysis of all adult and adolescent studies on reward outcome alterations. **b** Reward outcome alterations in adults with depression. **c** Reward outcome alterations in adolescents with depression. The right panel presents the results of the seed-based functional connectivity of the peak coordinates of altered reward outcome function in adults (putamen) and adolescents (mid cingulate cortex): **d** Resting-state functional connectivity of the caudate, **e** putamen and **f** mid cingulate cortex. The overlap of the putamen and MCC network maps is presented on the far right.

**Table 1.** Whole brain meta-analytic results of functional depression in adults and adolescents, significant at uncorrected p < 0.0025

| Meta-analysis | Contrast | Main brain regions | MNI coordinates | SDM Z | Voxels | I<sup>2</sup> statistic (%) | Egger's bias | Egger's p | Jackknife sensitivity |
| --- | --- | --- | --- | --- | --- | --- | --- | --- | --- |
| General | Depression < HC | R putamen/caudate | 12, 6, -8 | -4.213 | 316 | 4.46 | -0.38 | 0.638 | 31/30 |
|  | Depression > HC |  |  | None |  |  |  |  |  |
| Adults | Depression < HC | R putamen | 18, 4, -8 | -4.673 | 314 | 14.92 | -0.46 | 0.642 | 19/18 |
|  |  | Subgenual ACC | 0, 2, -10 | -3.832 | 38 | 2.58 | -0.15 | 0.880 |  |
|  | Depression > HC |  |  | None |  |  |  |  |  |
| Adolescents | Depression < HC | L median cingulate | 2, -28, 38 | -3.520 | 134 | 20.79 | -2.08 | 0.256 | 13/12 |
|  |  | R caudate nucleus | 12, -6, 16 | -3.276 | 16 | 4.04 | -0.79 | 0.663 |  |
|  |  | Inter-hemispheric | 2, 2, 8 | -3.117 | 12 | 11.70 | -0.92 | 0.643 |  |
|  | Depression > HC | R postcentral gyrus | 58, -14, 24 | 3.480 | 50 | 2.17 | -0.35 | 0.837 |  |

ACC, Anterior cingulate cortex, HC healthy controls, L, left, MNI, Montreal Neurological Institute, R right, SDM Seed-based D Mapping

### 3.3. Reward dysregulations on the network, behavioral, genetic and receptor levels

Respective comparative analyses were performed to characterize the identified regions on the network, behavioral, genetic and receptor levels. Network level analyses revealed the voxel-wise functional connectivity patterns of the common region (caudate) and peak separate regions in adults (putamen) and adolescents (MCC) respectively. The common region (caudate) connected more with the reward circuit pathways in the striatum as well as the frontal pole. The identified putamen region in adult depression coupled more with regions encompassing the bilateral dorsal striatal regions as well as lateral frontal extending to the insular regions. The MCC showed strong connectivity to frontal and midline structures resembling the default mode network (**Fig. 4a**). An overlap of the putamen and the MCC revealed strong connections to the bilateral frontal pole and bilateral occipital lobe.

**Fig. 4.**
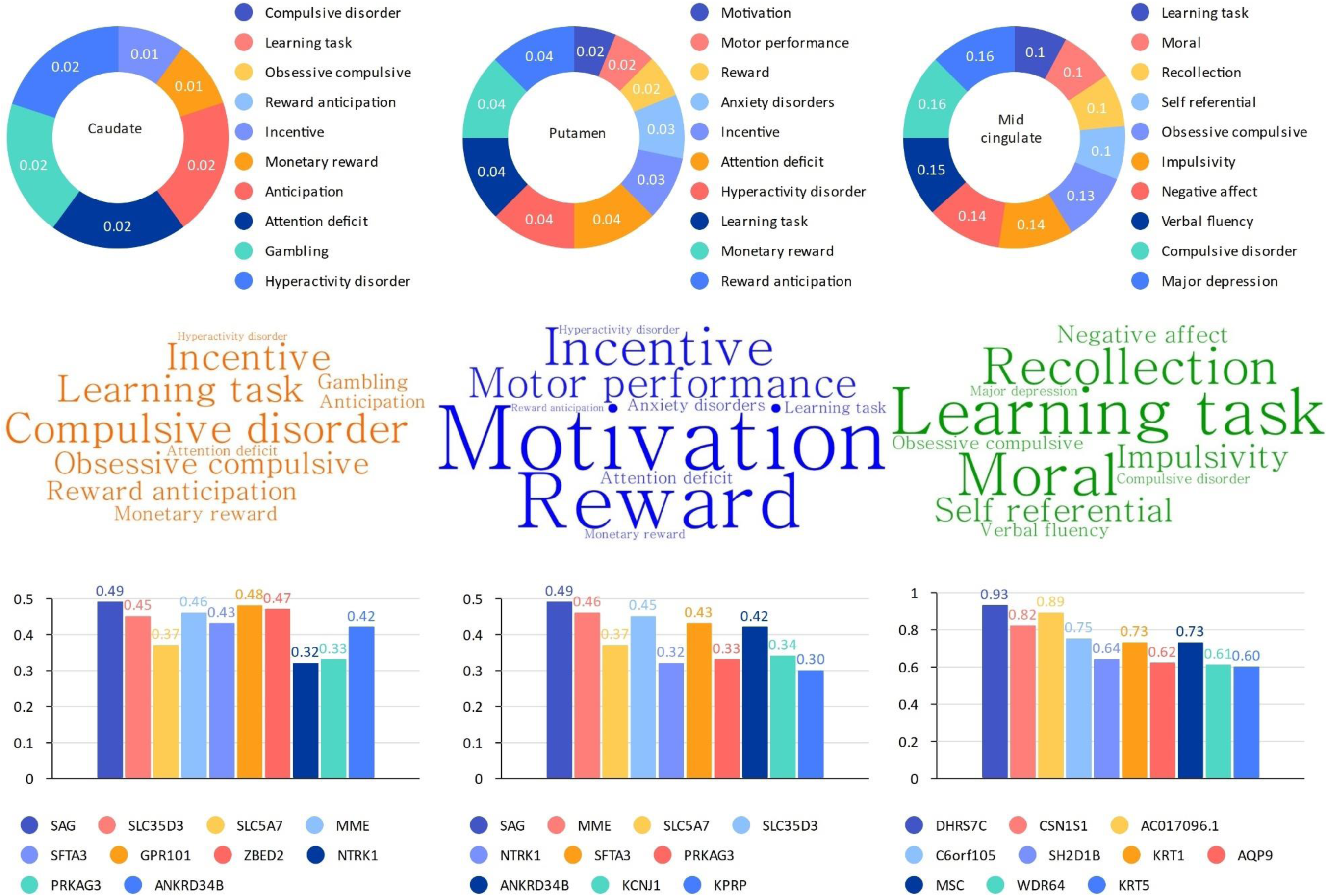
Results of the behavioral and genetic analyses of adult and adolescent depression. **a** Behavioral analysis of altered reward outcome in adults and adolescents (left), in adults only (middle) and adolescents (right). **b** A word cloud showing the behavioral terms. **c** Genetic level analysis showing the top ten genes identified in the key regions of caudate (left), putamen (middle) and mid cingulate (right).

Behavioral characterization of the identified regions revealed that the common region (caudate) was strongly associated with compulsive behaviors and reward. The putamen was strongly connected to motivation and behavioral control while the MCC coupled stronger with learning and social/moral cognition (**Fig. 4b**). In summary, behavioral characterization of adult and adolescent depression showed distinct behavioral patterns.

Genetic expression analyses revealed separable gene expressions for the adults and adolescents with depression. S-antigen visual arrestin (SAG) gene was the most expressed gene in the caudate and putamen. However, the DHRS7C (dehydrogenase 7C) was the most expressed gene in the MCC (**Fig. 4c**).

Receptor level analysis compared dopamine (DA) and serotonin (5HT) neurotransmitter mean densities in the peak regions. The results revealed that the putamen had a higher mean density of both neurotransmitters compared to the MCC. The mean density of DA and 5HT_1b_ in the putamen was 0.7074 and 0.4729 respectively (t=10.14, df=244, s.d=0.18). However, the mean density of DA and 5HT_1b_ in the mid cingulate cortex was 0.1632 and 0.1436 respectively (t=1.1452, df=244, s.d=0.13). The common region, caudate had a mean density of 0.4202 and 0.1171 for DA and 5HT_1b_ respectively (t=15.63, df=244, sd=0.21). The visual comparison of neurotransmitter expression in the putamen and the MCC is shown in the **Supplementary** Fig. 1.

### 3.4. Tests for heterogeneity, publication bias and sensitivity

Across all meta-analyses, the included studies exhibited low to moderate levels of heterogeneity. Tests for publication bias did not reach statistical significance, as indicated by bias p-values and symmetric funnel plots (**Supplementary** Fig. 2). The jackknife sensitivity analysis provided overall evidence supporting the robustness of our meta-analysis findings. A comprehensive interpretation of the results is available in **Table 1**.

### 3.5. Exploratory analyses

There were no significant regions of convergence in the adolescent VBM meta-analysis possibly due to limited number of studies. In line with our previous meta-analysis (Liu et al., 2022), the VBM meta-analysis in adults with depression revealed generally decreased gray matter volume in the right subgenual anterior cingulate (ACC), right middle temporal gyrus (MTG), right fusiform gyrus (FG) and left insula (**Supplementary** Fig. 3). Meta-regression in the fMRI data revealed that the mean age of adult patients was negatively associated with decreased reactivity in the putamen while a younger mean age of adolescents with depression was associated with a stronger reduction of MCC reactivity (**Supplementary** Fig. 4a and c). In addition, the number of adult female patients was positively associated with decreased activity in the putamen while the number of adolescent female patients was positively associated with decreased activity in the MCC (**Supplementary** Fig. 4b and d). Linear model analyses revealed that confounding effects of medication, age and gender showed no significant impact on the main meta-analytic results. Furthermore, subgroup meta-analyses of studies without medication in both adults and adolescents confirmed the stability of the main meta-analytic findings (**Supplementary** Fig. 4). Sub-analysis of unmedicated studies only, confirmed the stability of the findings (**Supplementary** Fig. 5).

## 4. Discussion

The current meta-analysis is the first to systematically examine common and age-specific neurofunctional alterations in depression compared to healthy controls during reward processing and revealed: a) Adults and adolescents with depression exhibit common reductions in subgenual ACC and caudate reward reactivity, b) Adolescents with depression specifically exhibit decreased reactivity in the mid cingulate cortex (MCC) and caudate nucleus, but increased activity in the postcentral gyrus while adults show reduced reactivity in the putamen and sugenual ACC, (c) the common striatal regions are characterized by reward and cognitive control processes and couple with dopaminergic reward circuits, while the MCC is characterized by social and learning processes and couples with the default mode network (DMN). All synthesized findings are considered according to the existing literature and implications for clinical practice.

### 4.1. Common alterations in the caudate

Consistent with our hypothesis, the caudate, which is a sub-region of the striatum, exhibited reduced neural reward sensitivity across adults and adolescents with depression. The caudate represents a highly-connected key reward processing hub with additional engagement in inhibitory and emotional processes (Oldham et al., 2018; Pizzagalli et al., 2009; Zhao et al., 2019; Zhuang et al., 2021). Previous neuroimaging meta-analyses have reported decreased caudate reward sensitivity in depression across reward domains (Bore et al., 2023; Zhang et al., 2013), and across patients over and below the age of 18 (Keren et al., 2018). Together with the present results and original studies reporting decreased caudal reward reactivity in children and adolescents with depression (Forbes et al., 2006; Forbes et al., 2009), the present findings underscore a key role of reduced caudate reward sensitivity as general markers of depression.

Further meta-analytic characterization suggests that dysfunction in this region lead to network level dysruptions in the fronto-striatal reward pathways, compulsive and reward alterations as well as alterations in S-antigen visual arrestin (SAG) gene signaling which is responsible for G-protein coupled receptor reactivity which mediates sensitivity to several neurotransmitters.

Overall, implications of the caudate in both adults and adolescents with depression suggests that the region critically mediates anhedonia and depressive episodes. Interventions targeting the caudate and associated functions could represent a general therapeutic strategy in depression.

### 4.2. Role and implications of the MCC in adolescent depression

Despite the extensive literature on striatal reward processing dysfunctions in depression, initial studies have suggested more widespread alterations in adolescent depression, including the insular, cingulate and frontal regions (Forbes et al., 2009; Gotlib et al., 2010). Our meta-analytic results indicated that adolescents with depression additionally exhibit decreased reward sensitivity in the MCC, the key hub for social behavior and social reward-based decision-making in the cingulate, supporting formation about the rewards that others will receive and the decisions that lead to other’s rewarding outcomes (Apps et al., 2013) or, the self-referential components of reward (Enzi et al., 2009).

Disruptions in these processes may be particularly impactful in adolescents navigating the complex social demands of their development. The behavioral and network characterization of the MCC further stressed its involvement in social and adaptive processes and communication with the DMN, a large-scale brain network strongly involved in social and self-referential cognition (Feng et al., 2021; Menon, 2023). Notably, the MCC also exhibited a higher serotonin density compared to the putamen (implicated in adult findings), suggesting potential differences in neurochemical underpinnings of adult and adolescent depression. Selective serotonin reuptake inhibitors (SSRIs) are first-line treatment for depression and potentially have more beneficial effects in childhood and adolescent depression (Mullen, 2018), suggesting that the serotonin system or interacting systems such as the oxytocin system (Lan et al., 2022) in combination with interventions targeting social functions, may have a high potential in adolescent depression.

### 4.3. Role and implications of the putamen in adult depression

In line with previous meta-analyses, our findings support the putamen as a neural substrate of reward processing disruptions in adult depression (Keren et al., 2018; Talati et al., 2022). Traditionally, the putamen has been associated with motor functions while recent studies showed more complex functions in other domains including reward processing and habit formation (Haber, 2016). Lower putamen activity is associated with greater anhedonia and individuals with smaller putamen volumes exhibited blunted responses during positive reward feedback (Auerbach et al., 2019; Schaub et al., 2021), which may link this region to anhedonia and habitual rather than reward-oriented behavior.

The exploratory VBM meta-analysis of adults with depression did not reveal neurostructural alterations in the identified striatal regions, while the findings resemble previous VBM meta-analysis reporting reduced gray matter in regions outside of the striatum in depression (Liu et al., 2022; Serra-Blasco et al., 2021), results may reflect a modifiable treatment target.

### 4.4 Limitations

Some limitations must be considered in the current meta-analysis. First, there was limited number of adolescent studies on reward anticipation to further segregate reward processing in both adults and adolescents. Second, the exploratory VBM studies on adolescents were only 7 and did not yield any significant clusters probably because of small sample size. Thirdly, some of the included studies had a mix of medicated and unmedicated patients in a single study.

Although the effects were considered in compiling the findings of the current study, medication effects could cause some errors if not properly controlled. It is suggested that future studies should include patients with identical medication status to limit heterogeneity and type II errors (Dichter et al., 2009).

## 5. Conclusions

Overall, altered reward processing is integral to the pathophysiology of depression. The current meta-analysis is the first to comprehensively reveal the neural correlates underlying dysfunctional reward processing in depression across the lifespan. In line with previous definitions, the caudate was identified as the region of common alterations in both adults and adolescents with depression while specific alterations of the putamen and mid cingulate cortex in adults and adolescents respectively, were unraveled. Distinct biomarkers representing age-specific alterations could serve as potential therapeutic interventions in current and future clinical practice.

### Declaration of Competing Interest

None reported.

## Funding

The present study was supported by the China Brain Project (MOST2030, Grant No. 2022ZD0208500), the National Natural Science Foundation of China (NSFC 82271583; 32250610208), and a startup grant from The University of Hong Kong.

## Role of the Funder/Sponsor

Any opinions, findings, conclusions, or recommendations expressed in this publication do not reflect the views of the Government of the Hong Kong Special Administrative Region or the Innovation and Technology Commission.

## Appendix A. Supplementary data

## Data Availability

Data supporting the findings of the present study is available on the Open Science Repository platform (OSF) (https://osf.io/v5nwj/).

## Notes

### Competing Interest Statement

The authors have declared no competing interest.

### Summary of Updates

Version includes additional technical specifications.

