## Supplementary Information for "Common and separable neural alterations in adult and adolescent depression – evidence from neuroimaging meta-analyses"

**Supplementary material**

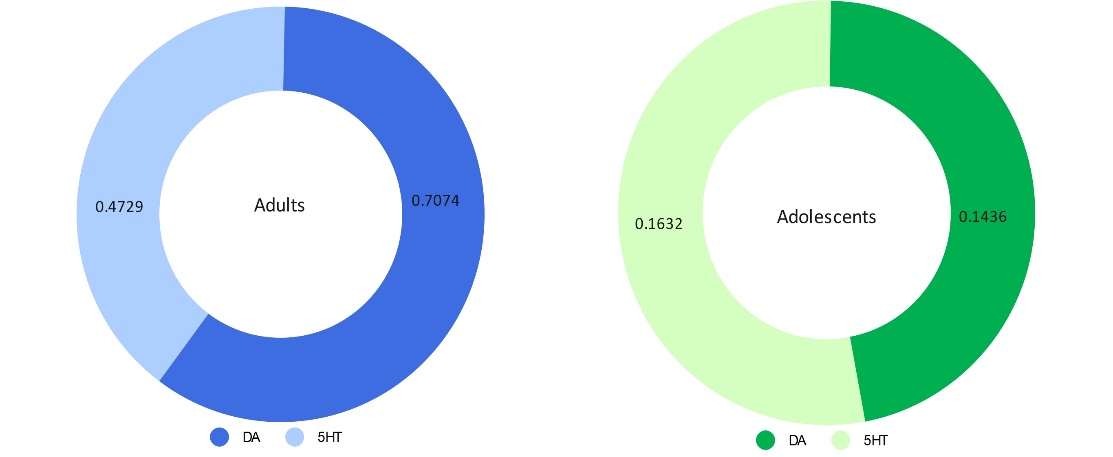

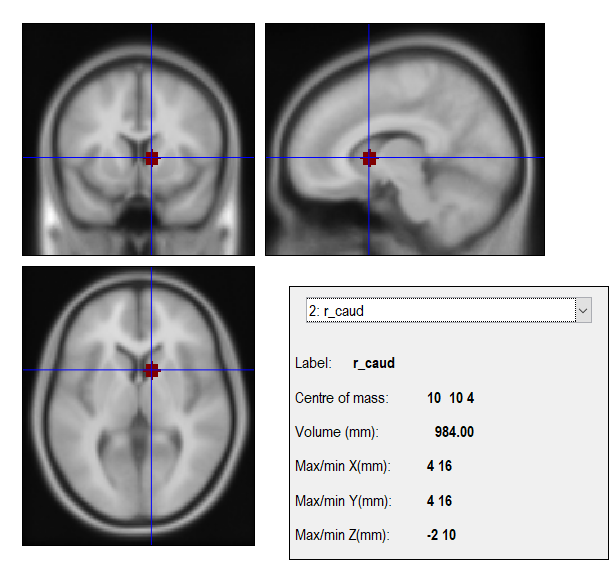

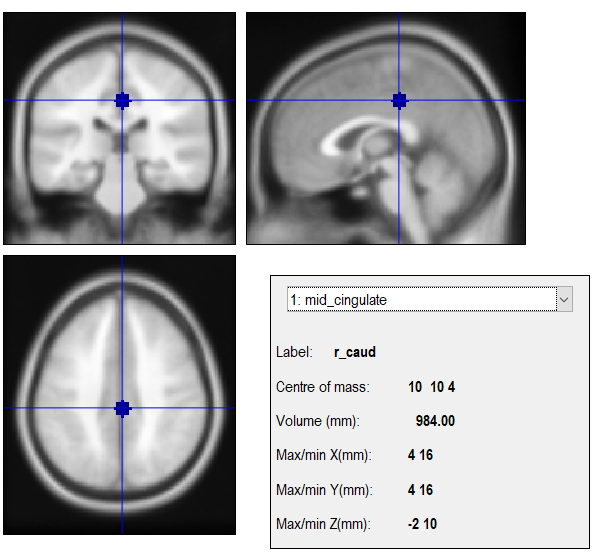

**Supplementary Fig. 1.** Receptor level characterization of DA and 5HT_1B_ in the right putamen (left) and MCC (right) respectively**.**

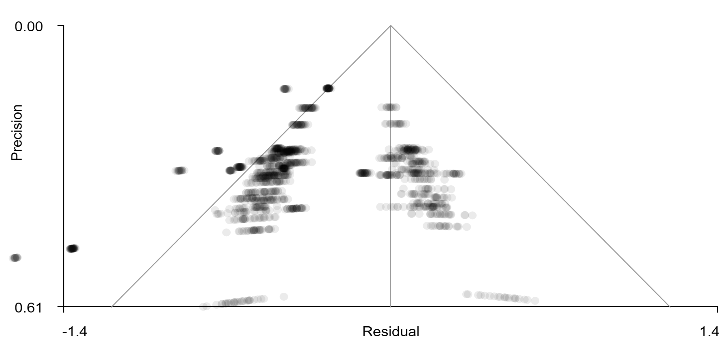

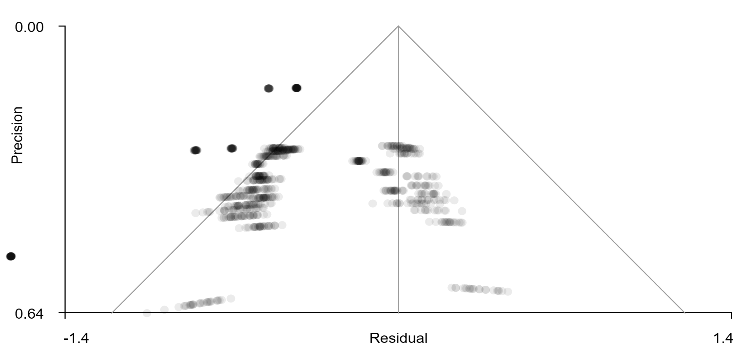

**a**

**b**

**c**

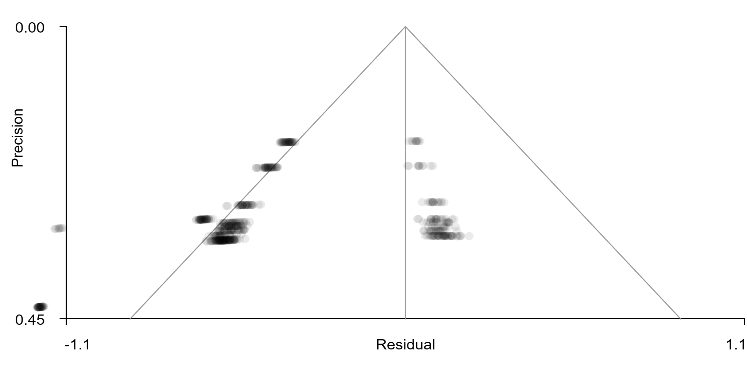

**Supplementary Fig. 2.** Funnel plots for the main clusters revealed in the main meta-analyses

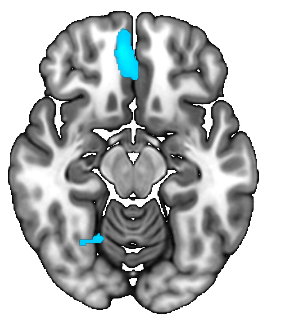

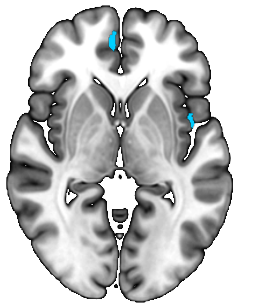

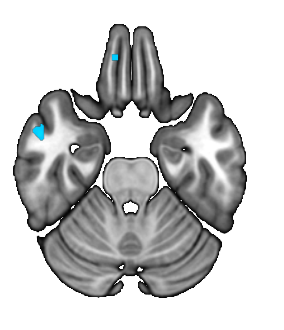

R FG

R sACC

R Insula

R MTG

**Supplementary Fig. 3.** Meta-analytic results of the exploratory VBM meta-analysis of adults with depression.

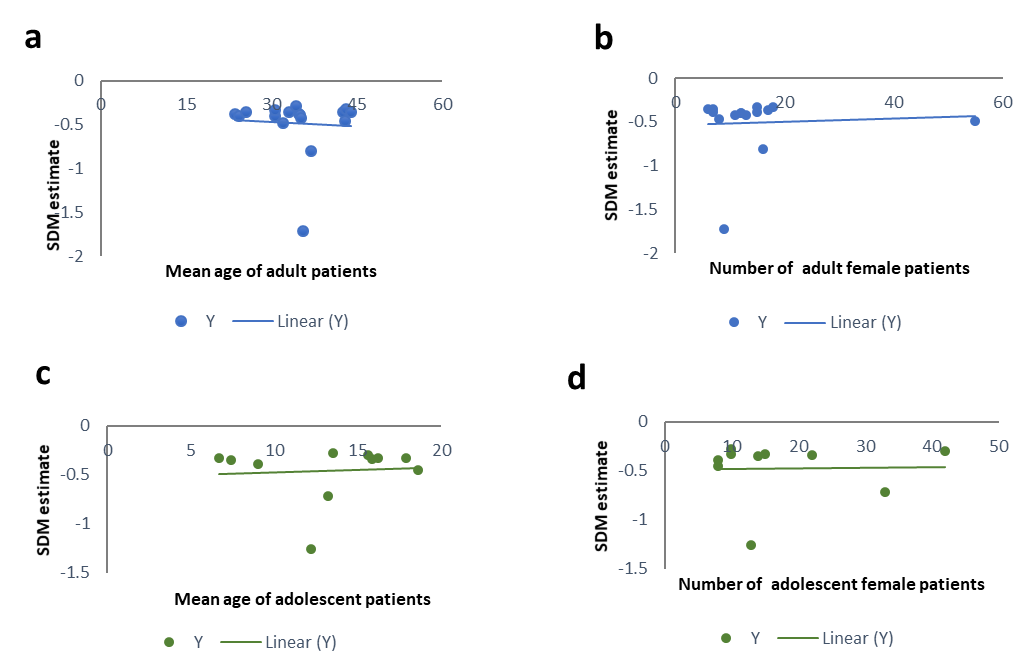

**Supplementary Fig. 4**. Regression analysis of age (a) and gender (b) in adults. Regression analysis of age (c) and gender (d) in adolescents.

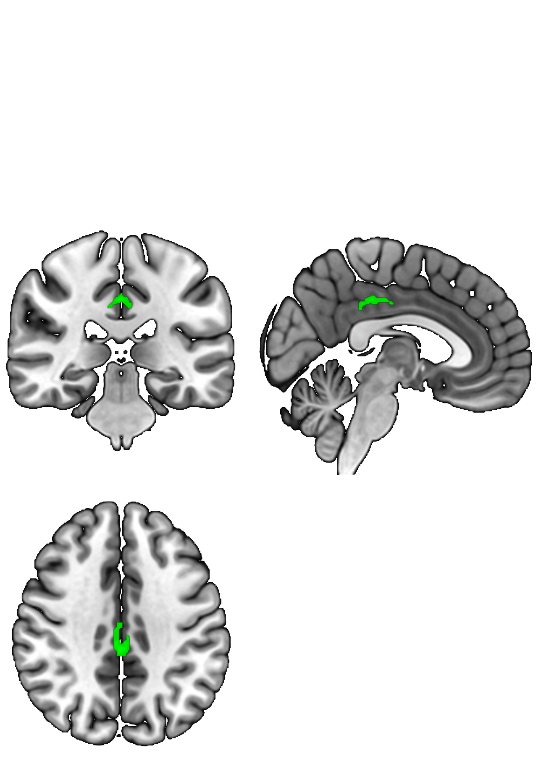

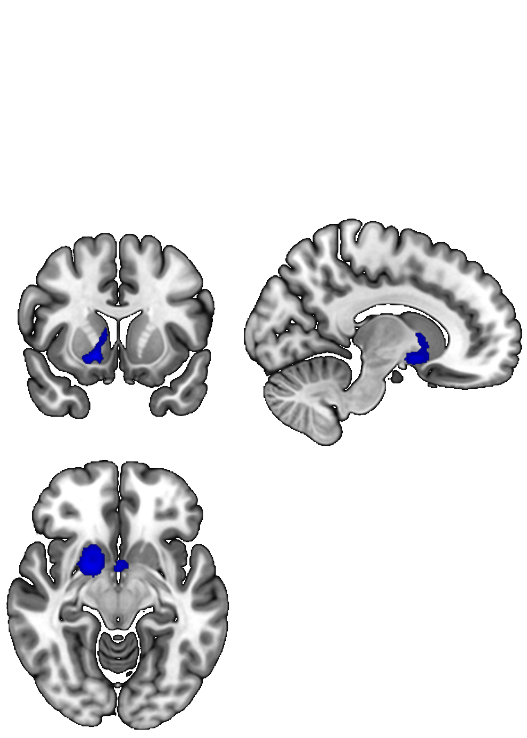

**a**

**b**

**Supplementary Fig. 5.** Meta-analysis results of unmedicated studies of adults (a) and adolescents (b).

**Supplementary Table 1**: PRISMA checklist 2020

| **Section and topic** | **Item #** | **Checklist item** | **Reported on page #** |
| --- | --- | --- | --- |
| **TITLE** | | | |
| Title | 1 | Identify the report as a systematic review. | 1 |
| **ABSTRACT** | | | |
| Abstract | 2 | See the PRISMA 2020 for Abstracts checklist | 3 |
| **INTRODUCTION** | | | |
| Rationale | 3 | Describe the rationale for the review in the context of existing knowledge. | 4,5,6 |
| Objectives | 4 | Provide an explicit statement of the objective(s) or question(s) the review addresses. | 6 |
| **METHODS** | | | |
| Eligibility criteria | 5 | Specify the inclusion and exclusion criteria for the review and how meta-analyses were grouped for the synthesis. | 8 |
| Information sources | 6 | Specify all databases, registers, websites, organisations, reference lists and other sources searched or consulted to identify studies. Specify the date when each source was last searched or consulted. | 7,8 |
| Search strategy | 7 | Present the full search strategies for all databases, registers and websites, including any filters and limits used. | 8 |
| Selection process | 8 | Specify the methods used to decide whether a meta-analysis met the inclusion criteria of the review, including how many reviewers screened each record and each report retrieved, whether they worked independently, and if applicable, details of automation tools used in the process. | 8 |
| Data collection process | 9 | Specify the methods used to collect data from reports, including how many reviewers collected data from each report, whether they worked independently, any processes for obtaining or confirming data from study investigators, and if applicable, details of automation tools used in the process | 8 |
| Data items | 10a | List and define all outcomes for which data were sought. Specify whether all results that were compatible with each outcome domain in each study were sought (e.g. for all measures, time points, analyses), and if not, the methods used to decide which results to collect. | - |
|  | 10b | List and define all other variables for which data were sought (e.g. participant and intervention characteristics, funding sources). Describe any assumptions made about any missing or unclear information. | Supplementary Table 2 |
| Meta-analysis quality assessment | 11 | Specify the methods used to assess quality in the included meta-analyses, including details of the tool(s) used, how many reviewers assessed each meta-analysis and whether they worked independently, and if applicable, details of automation tools used in the process | 7,8 |
| Effect measures | 12 | Specify for each outcome the effect measure(s) (e.g. risk ratio, mean difference) used in the synthesis or presentation of results. | 7,8 |
| Synthesis methods | 13a | Describe the processes used to decide which meta-analyses were eligible for each synthesis (e.g. tabulating the study intervention characteristics and comparing against the planned groups for each synthesis. | 8 Supplementary Table 2 |
|  | 13b | Describe any methods required to prepare the data for presentation or synthesis, such as handling of missing summary statistics, or data conversions. | 7 |
|  | 13c | Describe any methods used to tabulate or visually display results of individual meta-analyses. | 7 |
|  | 13d | Describe any methods used to synthesise results and provide a rationale for the choice(s). | 7 |
|  | 13e | Describe any methods used to explore possible causes of heterogeneity among study results (e.g. subgroup analysis, meta-regression). | 10 |
|  | 13f | Describe any sensitivity analyses conducted to assess robustness of the synthesised results | 10 |
| Reporting bias assessment | 14 | Describe any methods used to assess risk of bias due to missing results in a synthesis (arising from reporting biases). | 10 |
| Certainty assessment | 15 | Describe any methods used to assess certainty (or confidence) in the body of evidence for an outcome. | - |
| **RESULTS** | | |  |
| Study selection | 16a | Describe the results of the search and selection process, from the number of records identified in the search to the number of studies included in the review, ideally using a flow diagram. | Figure 1 |
|  | 16b | Cite studies that might appear to meet the inclusion criteria, but which were excluded, and explain why they were excluded. | Figure 1 |
| Study characteristics | 17 | Cite each included study and present its characteristics. | 8, Supplementary Table 2 |
| Quality of meta-analyses | 18 | Present assessments of quality for each included meta-analysis. | 13 |
| Results of individual meta-analyses | 19 | For all outcomes, present, for each meta-analysis: (a) summary statistics for each group (where appropriate) and (b) an effect estimate and its precision (e.g. confidence/credible interval), ideally using structured tables or plots. | 11,  Table 1 |
| Results of syntheses | 20a | For each synthesis, briefly summarise the characteristics and quality among contributing meta-analyses. | 11, Figure 3 |
|  | 20b | Present results of all statistical syntheses conducted. If meta-analysis was done, present for each the summary estimate and its precision (e.g. confidence/credible interval) and measures of statistical heterogeneity. If comparing groups, describe the direction of the effect. | 11, Table 1 |
|  | 20c | Present results of all investigations of possible causes of heterogeneity among study results. | 13, Table 1 |
|  | 20d | Present results of all sensitivity analyses conducted to assess the robustness of the synthesised results. | 13, Table 1 |
| Reporting biases | 21 | Present assessments of risk of bias due to missing results (arising from reporting biases) for each synthesis assessed. | 13, Table 1 |
| Certainty of evidence | 22 | Present assessments of certainty (or confidence) in the body of evidence for each outcome assessed. | - |
| **DISCUSSION** | | |  |
| Discussion | 23a | Provide a general interpretation of the results in the context of other evidence | 13 to 17 |
|  | 23b | Discuss any limitations of the evidence included in the review. | 13 to 17 |
|  | 23c | Discuss any limitations of the review processes used. | 13 to 17 |
|  | 23d | Discuss implications of the results for practice, policy, and future research | 13 to 17 |
| **OTHER INFORMATION** | | |  |
| Registration and protocol | 24a | Provide registration information for the review, including register name and registration number, or state that the review was not registered. | 7 |
|  | 24b | Indicate where the review protocol can be accessed, or state that a protocol was not prepared. | 7 |
|  | 24c | Describe and explain any amendments to information provided at registration or in the protocol. | - |
| Support | 25 | Describe sources of financial or non-financial support for the review, and the role of the funders or sponsors in the review. | 16 |
| Competing interests | 26 | Declare any competing interests of review authors. | 16 |
| Availability of data, code, and other materials | 27 | Report which of the following are publicly available and where they can be found: template data collection forms; data extracted from included studies; data used for all analyses; analytic code; any other materials used in the review. | 17 |

*From*: Page MJ et al., The PRISMA 2020 statement: an updated guideline for reporting systematic reviews. BMJ 2021;372. doi: 10.1136/bmj.n71. PRISMA 2020 has been originally designed for systematic reviews of individual studies.

**Supplementary Table 2. Demographic and Clinical characteristics of participants in the adolescent and adult depression**

| Study | No. of patients (f) | Mean age_P (s.d) | No. of HC (f) | Mean age_ HC (s.d) | Medication (%) | | Group | Reward-related task |
| --- | --- | --- | --- | --- | --- | --- | --- | --- |
| (Martin-Soelch et al., 2021) | 16 (12) | 24.31 (4.08) | 16 (12) | 25.19 (4.79) | 0 | Adults | | Monetary Incentive delay task |
| (Pizzagalli et al., 2009) | 26 (15) | 43.17 (12.98) | 31 (13) | 38.8 (14.48) | 0 | Adults | | Monetary Incentive delay task |
| (Dichter et al., 2012) | 19 (15) | 23.6 (4.09) | 19 (12) | 27.9 (6.3) | 0 | Adults | | Monetary Incentive delay task |
| (Hall et al., 2014) | 29 (16) | 37.01 (8.48) | 29 (16) | 37.38 (9.78) | 51.72 | Adults | | Reversal reward paradigm |
| (Smoski et al., 2009) | 14 (7) | 34.8 (14.3) | 15 (9) | 30.8 (9.70 | 0 | Adults | | Wheel of Fortune task |
| (Smoski et al., 2011) | 9 (NA) | 34.4 (15.1) | 13 (NA) | 26.2 (6.3) | 44.44 | Adults | | Monetary Incentive delay task |
| (Segarra et al., 2016) | 24 (7) | 33.08 (9.15) | 21 (4) | 34.33 (10.11) | 54.16 | Adults | | Simulated slot-machine game |
| (Admon et al., 2015) | 26 (NA) | 42.66 (11.72) | 29 (NA) | 37.75 (14.05) | 0 | Adults | | Monetary Incentive delay task |
| (Knutson et al., 2008) | 14 (NA) | 30.71 (8.8) | 12 (NA) | 28.67 (4.25) | 0 | Adults | | Monetary Incentive delay task |
| (Oh et al., 2021) | 108 (55) | 31.98 (13.09) | 108 (86) | 29.86 (10.58) | 0 | Adults | | Classic juice-delivery |
| (Epstein et al., 2006) | 10 (9) | 35.6 (NA) | 12 (7) | 32 (NA) | 0 | Adults | | Paradigm involving 3- word types |
| (Gradin et al., 2015) | 25 (17) | 24.48 (5.52) | 25 (17) | 25.44 (5.02) | 0 | Adults | | Social fairness using ultimatum game |
| (Keedwell et al., 2005) | 12 (8) | 43 (9.8) | 12 (8) | 36 (14.6) | 91.67 | Adults | | Happy and Sad emotional stimuli |
| (McCabe et al., 2009) | 13 (11) | NA | 14 (9) | NA | 46.15 | Adults | | Sight and taste of chocolate or moldy strawberries |
| (Kumari et al., 2003) | 6 (6) | 44 (NA) | 6 (6) | 43.5 (NA) | 0 | Adults | | Paradigm of evoking affect with picture-caption pairs |
| (Canli et al., 2004) | 15 (12) | 35.1 (NA) | 15 (12) | 30.7 (NA) | 46.7 | Adults | | Lexical decision task |
| (Fournier et al., 2013) | 26 (18) | 30.6 (7.8) | 28 (16) | 32.6 (3.4) | 26.92 | Adults | | Emotional dynamic face task |
| (Gotlib et al., 2005) | 18 (13) | 35.2 (10.26) | 18 (13) | 30.8 (NA) | 50 | Adults | | Pictures of emotional faces |
| (Fischer et al., 2019) | 15 (NA) | 17.87 (2.68) | 18 (NA) | 19.09 (2.93) | 0 | Adolescents | | Event-related MIDT |
| (Gotlib et al., 2010) | 13 (13) | 12.2 (1.7) | 13 (13) | 12.7 (1.4) | 0 | Adolescents | | Social Incentive delay task |
| (Olino et al., 2015) | 17 (NA) | NA | 23 (NA) | NA | 0 | Adolescents | | Card guessing |
| (Morgan et al., 2019) | 25 (15) | 6.72 (NA) | 31 (16) | 6.87 (NA) | 0 | Adolescents | | Response to happy faces |
| (Forbes et al., 2009) | 15 (10) | 13.5 (2.1) | 28 (21) | 13.1 (2.6) | 0 | Adolescents | | Card guessing |
| (Sharp et al., 2014) | 33 (33) | 13.21 (1.85) | 19 (19) | 13.71 (1.85) | 0 | Adolescents | | Card guessing |
| (Chantiluke et al., 2012) | 20 (10) | 16.2 (0.8) | 21 (11) | 16.3 (1.1) | 0 | Adolescents | | Monetary task |
| (Wiggins et al., 2017) | 27 (14) | 7.44 (0.73) | 19 (14) | 7.64 (0.84) | 0 | Adolescents | | Monetary Incentive delay task |
| (Davey et al., 2011) | 19 (11) | 18.6 (2.2) | 20 (12) | 19.3 (2.9) | 52.9 | Adolescents | | Positive-feedback faces |
| (Luking et al., 2016) | 16 (8) | 9.05 (1.14) | 32 (17) | 9.28 (1) | 0 | Adolescents | | Card guessing |
| (Harlé et al., 2023) | 61 (42) | 15.6 (1.4) | 66 (33) | 15.3 (1.4) | 0 | Adolescents | | Reward-seeking paradigm |
| (Yoon et al., 2022) | 45 (23) | 15.88 (1.05) | 43 (19) | 15.43 (1.18) | 0 | Adolescents | | Gambling task |

F, females, MIDT monetary incentive delay task, N number, NA not available, P patients, SD standard deviation, SIDT social incentive delay task

**Supplementary Table 3.** Summary of the VBM studies included

| **Study** | **No. of patients** | **No. of HC** | **Age_P (s.d)** | **Age_HC (s.d)** | **Medication (%)** | **Category** |
| --- | --- | --- | --- | --- | --- | --- |
| (Straub et al., 2019) | 60 | 43 | 17.3 (3.44) | 17.62 (3.85) | 55 | Adolescents |
| (Shad et al., 2012) | 22 | 22 | 15 (2.1) | 16 (2.1) | 18 | Adolescents |
| (Li et al., 2015) | 42 | 30 | 20.26 (0.89) | 20.2 (1.3) | 0 | Adolescents |
| (Vulser et al., 2015) | 119 | 461 | 14.45 (0.36) | 14.4 (0.41) | 0 | Adolescents |
| (Wehry et al., 2015) | 14 | 41 | 14 (3) | 13 (2) | NA | Adolescents |
| (Zhang et al., 2012) | 33 | 32 | 20.52 (1.72) | 21.03 (1.47) | 0 | Adolescents |
| (Pannekoek et al., 2014) | 26 | 26 | 15.4 (1.5) | 14.7 (1.5) | 0 | Adolescents |
| (Wang et al., 2020) | 36 | 27 | 34.11 (10.39) | 32.44 (8.57) | 0 | Adults |
| (Zhang et al., 2021) | 20 | 20 | 28 (9.1) | 31.7 (11.4) | 0 | Adults |
| (van de Mortel et al., 2022) | 88 | 27 | 50.77 (12) | 60.7 (8) | 100 | Adults |
| (Li et al., 2017) | 23 | 25 | 49.91 (8.44) | 49.2 (10.25) | 0 | Adults |
| (Zhang et al., 2020) | 30 | 63 | 25 (NA) | 23 (NA) | 0 | Adults |
| (Yang et al., 2017) | 82 | 82 | 28.84 (9.17) | 27.72 (8) | 0 | Adults |
| (Meng et al., 2020) | 159 | 53 | 33.71 (10.21) | 35.64 (8.66) | 0 | Adults |
| (Romanczuk-Seiferth et al., 2014) | 63 | 63 | 25 (NA) | 26 (NA) | 0 | Adults |
| (Liu et al., 2012) | 17 | 17 | 26.71 (7.73) | 24.24 (4.41) | 100 | Adults |
| (Lan et al., 2016) | 27 | 27 | 41.5 (16) | 39.2 (16.7) | NA | Adults |
| (Brosch et al., 2022) | 110 | 110 | 38.35 (11.7) | 38.83 (12.21) | 71 | Adults |
| (Depping et al., 2015) | 22 | 22 | 33.5 (8.9) | 31.4 (11.2) | 100 | Adults |
| (Guo et al., 2014) | 44 | 44 | 27.52 (8.57) | 29.39 (6.7) | 0 | Adults |
| (Jung et al., 2014) | 24 | 29 | 43 (10.1) | 43.6 (13.4) | 100 | Adults |
| (Kong et al., 2014) | 28 | 28 | 34.42 (8.24) | 32.07 (9.27) | 0 | Adults |
| (Lai and Wu, 2014) | 38 | 27 | 36.57 (5.46) | 38.29 (11.8) | 0 | Adults |
| (Lai and Wu, 2015) | 53 | 54 | 40.07 (8.99) | 40.38 (10.51) | 0 | Adults |
| (Lai et al., 2016) | 15 | 27 | 37.46 (5.93) | 38.29 (11.8) | 100 | Adults |
| (Liu et al., 2022) | 64 | 61 | 28.47 (9.74) | 30.49 (9.55) | 0 | Adults |
| (Mak et al., 2009) | 17 | 17 | 45.5 (8.5) | 45.8 (9.8) | 100 | Adults |
| (Nan et al., 2020) | 166 | 166 | 33.69 (9.99) | 34.64 (10.98) | 0 | Adults |
| (Opel et al., 2016) | 20 | 20 | 37.9 (10.9) | 36.3 (12.1) | 0 | Adults |
| (Ozalay et al., 2016) | 24 | 24 | 46.2 (3.9) | 47.3 (5.6) | 0 | Adults |
| (Peng et al., 2011) | 22 | 30 | 46.7 (8.9) | 45.9 (9) | 22 | Adults |
| (Xu et al., 2019) | 11 | 12 | 39.27 (7.84) | 39.08 (7.4) | 0 | Adults |
| (Zhuo et al., 2017) | 45 | 48 | 38.8 (13.3) | 38.6 (10.5) | 91 | Adults |
| (Chen et al., 2018) | 79 | 47 | 30.7 (8.5) | 29.7 (9.2) | 0 | Adults |
| (Abe et al., 2010) | 21 | 42 | 48.1 (13.5) | 48 (13.2) | 90 | Adults |
| (Grieve et al., 2013) | 102 | 34 | 33.8 (13.1) | 31.5 (12.4) | 0 | Adults |
| (Kandilarova et al., 2019) | 39 | 42 | 47.7 (13.9) | 42.6 (13.7) | 95 | Adults |
| (Klauser et al., 2015) | 56 | 33 | 34.02 (8.96) | 34.71 (9.93) | 59 | Adults |
| (Lee et al., 2011) | 47 | 51 | 46 (9.1) | 45.7 (8) | 64 | Adults |
| (Lu et al., 2018) | 76 | 86 | 34.27 (8.37) | 33.35 (7.62) | 0 | Adults |
| (Lu et al., 2019) | 30 | 48 | 23.98 (5.25) | 21.5 (3.84) | 0 | Adults |
| (Nakano et al., 2014) | 36 | 54 | 49 (11.4) | 45.4 (16.1) | 86 | Adults |
| (Salvadore et al., 2011) | 58 | 107 | 38.8 (11.1) | 36.2 (10.3) | 75 | Adults |
| (Ueda et al., 2016) | 30 | 48 | 44.3 (13) | 41.2 (11.4) | 0 | Adults |
| (Wagner et al., 2011) | 30 | 30 | 37.55 (11.5) | 35.1 (10.4) | 0 | Adults |
| (Chen et al., 2016) | 27 | 28 | 33 (10.8) | 33 (11.7) | 0 | Adults |
| (Bergouignan et al., 2009) | 20 | 21 | 33.16 (9.58) | 28.21 (5.5) | 100 | Adults |
| (Scheuerecker et al., 2010) | 13 | 15 | 37.9 (10.1) | 35.5 (10.9) | 0 | Adults |
| (Arnone et al., 2013) | 39 | 66 | 36.3 (8.8) | 32.1 (9.3) | 0 | Adults |
| (Sprengelmeyer et al., 2011) | 17 | 21 | 45.6 (12.3) | 42 (12.9) | 100 | Adults |

F, females, H, healthy controls, NA, not available, No. number, P, patients, s.d, standard deviation

**Supplementary Table 4.** Short summary of the top 10 genes expressed in the caudate, putamen, and MCC. The corresponding characterizations were acquired from PubMed gene.

| **GENE** | **SUMMARY** |
| --- | --- |
| SAG (S-antigen visual arrestin) | An Agonist-mediated desensitization of G-protein-coupled receptors and cause inhibit some cellular responses to neurotransmitters, hormones, or sensory signals. |
| MME (Membrane metalloendopeptidase) | Inactivates some peptide hormones such as oxytocin, substance P, glucagon, enkephalins, bradykinin and neurotensin. |
| SLC5A7 (Solute carrier family 5 member 7) | When disrupted, it results in implications of disorders such as depression, schizophrenia and attention-deficit disorder. |
| SLC35D3 (Solute carrier family 35-member D3) | Carries carbohydrate and pyrimidine nucleotide-sugar. |
| NTRK1 (neurotrophic receptor tyrosine kinase 1) | Kinase present in the gene facilitates cell differentiation and identifies sensory neuron subtypes. |
| SFTA3 (Surfactant associated 3) | Involved in healing wounds. |
| GPR101 (G protein-coupled receptor 101) | The protein encoded by this gene is less known. |
| ZBED2 (zinc finger BED-type containing 2) | Predicted to be in the nucleus and is likely embedded in chromatin. |
| PRKAG3 (Protein kinase AMP-activated non-catalytic subunit gamma 3) | AMPK is a crucial enzyme that observes the energy status of cells as well as regulating the body’s metabolism. |
| ANKRD34B (ankyrin repeat domain 34B) | Predictions show that the gene is found with cells (cytoplasm and nucleus). |
| KCNJ1 (potassium inwardly rectifying channel subfamily J member 1) | It is stimulated by internal ATP. It plays a key role of balancing potassium in the body. |
| KPRP (keratinocyte proline rich protein) | The gene enables differentiation of keratinocyte. |
| DHRS7C (dehydrogenase/reductase 7C) | Facilitates activity of NAD-retinol dehydrogenase as well as regulating release of calcium ion. |
| CSN1S1 (casein alpha s1) | Predictions show involvement in estradiol, dehydroepiandrosterone and steroid hormone responses. |
| C6orf105 | Involved in hydrolase activity. |
| SH2D1B (SH2 domain containing 1B) | Regulates signal transduction using expressed receptors that lie on the surface of cells. |
| KRT1 (keratin 1) | Expressed through differentiation of epithelial tissues. |
| AQP9 (aquaporin 9) | Encodes aquaporins which facilitates transportation of urea and osmotic water. |
| MSC (musculin) | Target for the signal transduction pathway of the B-cell receptor. |
| KRT5 (keratin 1) | Mutations in this gene has been associated with epidermolysis bullosa simplex. |
| WDR64 (WD repeat domain 64) | N/A |
| AC017096.1 | N/A |
